# Focal Cerebral Arteriopathy Severity Score Validation and Temporal Dynamics in Korean Pediatric Stroke: Distinguishing Inflammatory Arteriopathy from Unilateral Moyamoya Disease

**DOI:** 10.64898/2025.12.03.25341601

**Authors:** Seungjae Lee, Woo Joong Kim, Soo Yeon Kim, Seungbok Lee, Ji Yeoun Lee, Ji Hoon Phi, Seung-Ki Kim, Jong-Hee Chae, Byungchan Lim

**Author notes:** Correspondence: Byungchan Lim, MD, PhD, Department of Pediatrics, Seoul National University Children’s Hospital, Seoul National University College of Medicine, Jongro-Gu, Daehak-ro 101, Seoul, Republic of Korea.

## Abstract

**BACKGROUND AND PURPOSE:** Focal cerebral arteriopathy-inflammatory type (FCA-i) is a leading cause of pediatric arterial ischemic stroke, but diagnostic challenges persist, particularly in East Asian populations where moyamoya disease (MMD) prevalence is high. The Focal Cerebral Arteriopathy Severity Score (FCASS) quantifies arteriopathy severity but has not been validated in East Asian cohorts. We aimed to validate FCASS in Korean pediatric patients with FCA-i and compare temporal progression patterns with unilateral moyamoya disease.

**METHODS:** We conducted a retrospective cohort study of children with arterial ischemic stroke presenting to Seoul National University Hospital between January 2002 and December 2024. Patients were classified according to Childhood Arterial Ischemic Stroke Standardized Classification and Diagnostic Evaluation criteria. The FCASS was applied to serial magnetic resonance angiograms at baseline, peak severity, and final follow-up. Clinical outcomes were assessed using the Pediatric Stroke Outcome Measure at 12 months.

**RESULTS:** Among 216 children with arterial ischemic stroke, 132 patients (61.1%) demonstrated arteriopathy, including 49 with FCA-i and 60 with MMD. In FCA-i patients, the severity score correlated significantly with baseline infarct burden (ρ=0.42, *P*=0.0069) and exhibited characteristic monophasic evolution with early peak at 2 months followed by gradual recovery reaching lowest values at 11 months. Unilateral MMDpatients demonstrated consistently higher severity scores at all timepoints compared with FCA-i (baseline: 6.0 vs 2.0; final: 8.0 vs 3.0, *P*<0.001) without radiographic recovery. A baseline severity score ≥8.0 predicted contralateral progression in unilateral MMD with area under the curve of 0.962 (sensitivity 0.83, specificity 0.91).

**CONCLUSIONS:** The FCASS demonstrates validity as a dynamic biomarker for monitoring FCA-i in Korean pediatric patients, exhibiting characteristic monophasic recovery patterns that distinguish it from progressive unilateral MMD.

---

Pediatric arterial ischemic stroke (AIS) is uncommon, with an estimated annual incidence of 4 to 13 per 100,000 children, and represents a leading cause of long-term neurological morbidity in childhood.^1,2^ The etiologic spectrum of pediatric AIS differs markedly from that of adults, with non-atherosclerotic mechanisms predominating. Among the various risk factors, intracranial arteriopathies have emerged as the most frequent cause, accounting for more than half of cases and serving as the strongest predictor of stroke recurrence and poor outcomes.^3–5^

Focal cerebral arteriopathy (FCA) is a key subtype of pediatric arteriopathy that has garnered increasing attention over the past two decades as a leading cause of AIS in previously healthy children.^6^ Initially described as “Transient Cerebral Arteriopathy” due to its monophasic and often reversible course, FCA is now recognized as a heterogeneous entity encompassing multiple subtypes, including inflammatory (FCA-i), dissection-related (FCA-d), and undefined forms.^4,7,8^

The inflammatory subtype, FCA-i, is the most frequent form and is presumed to result from a post-infectious immune-mediated process.^4,9^ It is characterized angiographically by unilateral, unifocal stenosis or irregularity of large intracranial arteries in the anterior circulation, commonly involving the distal internal carotid artery (ICA), M1, and A1 segments.^4,10^ Clinical presentation typically includes acute-onset hemiparesis or focal neurological deficits in school-aged children, with infarcts localized to the basal ganglia or deep middle cerebral artery territory.^10,11^

Diagnostically, FCA poses significant challenges. While magnetic resonance angiography (MRA) serves as the standard vascular imaging modality, its sensitivity remains limited, and definitive diagnosis often requires serial imaging to distinguish FCA from mimics such as intracranial dissection, primary central nervous system (CNS) vasculitis, and early moyamoya disease (MMD).^12,13^ The Childhood AIS Standardized Classification and Diagnostic Evaluation (CASCADE) criteria was developed to improve diagnostic reliability across pediatric stroke etiologies. However, inter-rater agreement remains lowest for FCA (κ=0.49), highlighting persistent ambiguity in its clinical and radiographic definition.^14^

To address the need for standardized severity assessment, Fullerton et al. developed the Focal Cerebral Arteriopathy Severity Score (FCASS), which quantifies stenosis across five anterior circulation segments.^4,15^ FCASS demonstrates significant correlation with infarct volume and 12-month neurological outcomes and has been independently validated in Swiss and North American cohorts.^15–17^

Unilateral MMD further complicates this diagnostic landscape. While confirmed MMD requires bilateral terminal ICA or proximal Circle of Willis stenosis with moyamoya collaterals, unilateral presentations have been increasingly recognized, accounting for approximately 10%–18% of pediatric MMD cases.^7,18,19^ The natural history of unilateral MMD is heterogeneous such that up to half of affected children progress to bilateral disease on long-term follow-up, whereas others remain stable or even demonstrate improvement.^19–21^ These uncertainties make early differentiation from monophasic arteriopathies such as FCA-i especially challenging, emphasizing the need to better define its clinical course and management.

Despite these advances, important knowledge gaps persist regarding the natural history and optimal management of FCA-i, particularly in diverse populations.^7,10^ Most existing literature derives from North American and European cohorts, with limited data from East Asia where the prevalence of MMD is significantly higher. In this region, distinguishing FCA-i from early-stage unilateral MMD presents challenges due to overlapping clinical and radiological features, especially in the anterior circulation.^18^

The aim of the present study is to characterize FCA-i in a Korean pediatric cohort using the FCASS framework and to compare its temporal progression and severity with unilateral MMD. Through retrospective serial imaging analysis, we sought to delineate distinguishing features among these entities and propose evidence-based strategies for early diagnosis and follow-up imaging protocols tailored to populations where both conditions are prevalent.

## MATERIALS AND METHODS

### Study Design and Patient Selection

We conducted a retrospective cohort study at Seoul National University Children’s Hospital, a tertiary academic center in Korea. Children aged 1 month to 18 years diagnosed with acute AIS between January 1, 2002, and December 31, 2024, were identified through institutional electronic medical records using ICD-9 codes 433.01–435.9 and 768.01–768.9, and ICD-10 codes I63.00–I63.9 and P91.0. Patients with perinatal stroke, hemorrhagic stroke, venous infarction, or diffuse hypoxic injury were excluded. The study was approved by Seoul National Hospital’s institutional review board, which waived the need for written informed consent based on minimal risk (IRB No. H-2508-155-1670). Stroke subtype classification followed the CASCADE criteria, with patients categorized into arteriopathic and non-arteriopathic groups and further subclassified into FCA, dissection, MMD, and vasculitis based on imaging review and clinical correlation.^4,13^

### Arteriopathy Definitions

FCA-i was defined as unilateral, unifocal stenosis or irregularity of large intracranial arteries in the anterior circulation without dissection or moyamoya collaterals, consistent with presumed inflammatory etiology.^4,12^ Diagnosis was supported by monophasic course, recent infection history, and vessel wall imaging findings when available. Unilateral MMD was defined as steno-occlusive disease of the terminal ICA and proximal branches with basal collateral formation, without contralateral involvement at baseline.^7,22^

### Neuroimaging Protocol and Severity Assessment

All patients underwent brain MRI and time-of-flight MRA using 1.5T or 3.0T scanners. Imaging studies were retrospectively reviewed by a pediatric neurology team blinded to clinical outcomes. The FCASS was applied to serial MRAs to quantify arteriopathy severity.^15^ Five arterial segments on the affected side (supraclinoid ICA, M1, M2, A1, A2) were scored from 0 to 4 based on the severity criteria as follows: 0 = no involvement; 1 = irregularity or banding with no stenosis; 2 = stenosis with less than 50% reduction in diameter; 3 = stenosis with more than 50% reduction in diameter; 4 = occlusion. Any involvement of the A2 or M2 segments was assigned a score of 3 to account for the vessel caliber. FCASS was calculated at baseline, worst point, and final imaging. For patients with serial MRAs, changes were assessed across predefined intervals (0–3, 3–6, 6–12, and 12–24 months) to enable time-series evaluation of progression and recovery. To confirm final arteriopathy classification, patients were required to have at least one follow-up MRA obtained 6–12 months after onset, unless death or loss to follow-up occurred.

### Clinical Data Collection

Clinical variables included demographics, presenting symptoms (hemiparesis, facial palsy), and recent infection history within 12 months, including varicella zoster virus. Treatment was categorized as antiplatelet monotherapy, combination antithrombotic therapy, or corticosteroid use. Neurological outcomes were assessed using the Pediatric Stroke Outcome Measure (PSOM) at approximately 12 months.^23^ Baseline infarct burden was quantified using the modified pediatric Alberta Stroke Program Early CT Score (modASPECTS) on diffusion-weighted imaging.^24^

### Statistical Analysis

Continuous variables are reported as median with interquartile ranges, and categorical variables as counts and percentages. Between-group comparisons were performed using the Mann–Whitney *U* test for continuous variables and Fisher exact test for categorical variables. Longitudinal FCASS changes were analyzed using Wilcoxon signed-rank tests, with temporal intervals compared using Bonferroni-adjusted pairwise comparisons. Correlation between modASPECTS and baseline FCASS was assessed using Spearman rank correlation. Receiver operating characteristic analysis evaluated the discriminatory value of baseline FCASS for identifying progressive disease. All analyses were performed using R version 4.3.0 (R Foundation for Statistical Computing), with two-tailed *P*<0.05 considered statistically significant.

## RESULTS

### Cohort Characteristics and Arteriopathy Classification

Between January 2002 and December 2024, 216 children with AIS were identified through standardized ICD-9/10 codes and electronic medical record review. Of these, 132 patients (61.1%) were classified as having arteriopathy, while 84 (38.9%) showed no vascular abnormality. Arteriopathies were subclassified as FCA-i(n=49), MMD (n=60), arterial dissection (n=13), and other less common causes.

### Clinical Characteristics of Arteriopathy Subtypes

Among 49 FCA-i cases, 41 (84%) involved the anterior circulation, with a median age of 8.6 years (IQR, 6.4–11.3) and male predominance (55%). Hemiparesis was the most common presenting symptom (88%), followed by facial palsy (29%) and aphasia (24%). Only 4% had documented varicella zoster virus infection within 12 months. Anti-inflammatory therapies, including corticosteroids or intravenous immunoglobulin, were administered in 12% of patients, while dual antithrombotic therapy was used in 33%. Neurological deficits at discharge were observed in 65% of patients, but 12-month PSOM scores demonstrated favorable outcomes (median 0.5; IQR, 0.0–1.0). No deaths occurred, and stroke recurrence was rare (4%).

In contrast, 60 patients with MMD presented at a younger median age of 5.7 years (IQR, 3.1–9.2), with a female predominance (57%). Hemiparesis was the leading symptom (90%), followed by dysarthria (22%) and facial palsy (18%). Almost all patients (98%) underwent revascularization surgery.

Finally, 13 patients with arterial dissection had a median age of 7.0 years (IQR, 3.1–10.4) and male predominance (62%). Hemiparesis was also the most frequent initial presentation (61%), and facial palsy was reported in 15%. A preceding history of head or neck trauma was identified in 38%. Most patients received combined antithrombotic therapy (54%), and 15% underwent revascularization surgery (Table 1).

**Table 1.**
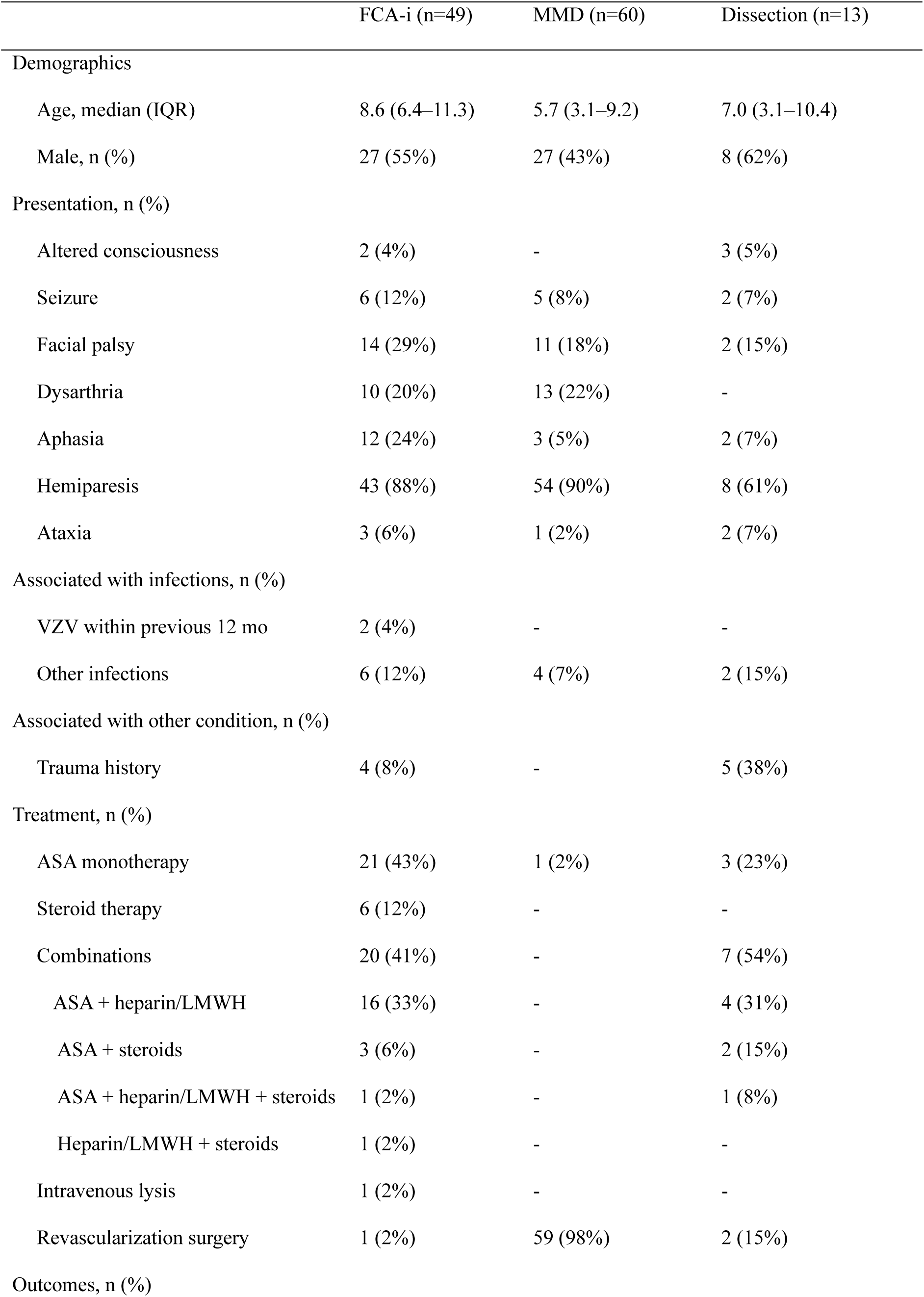
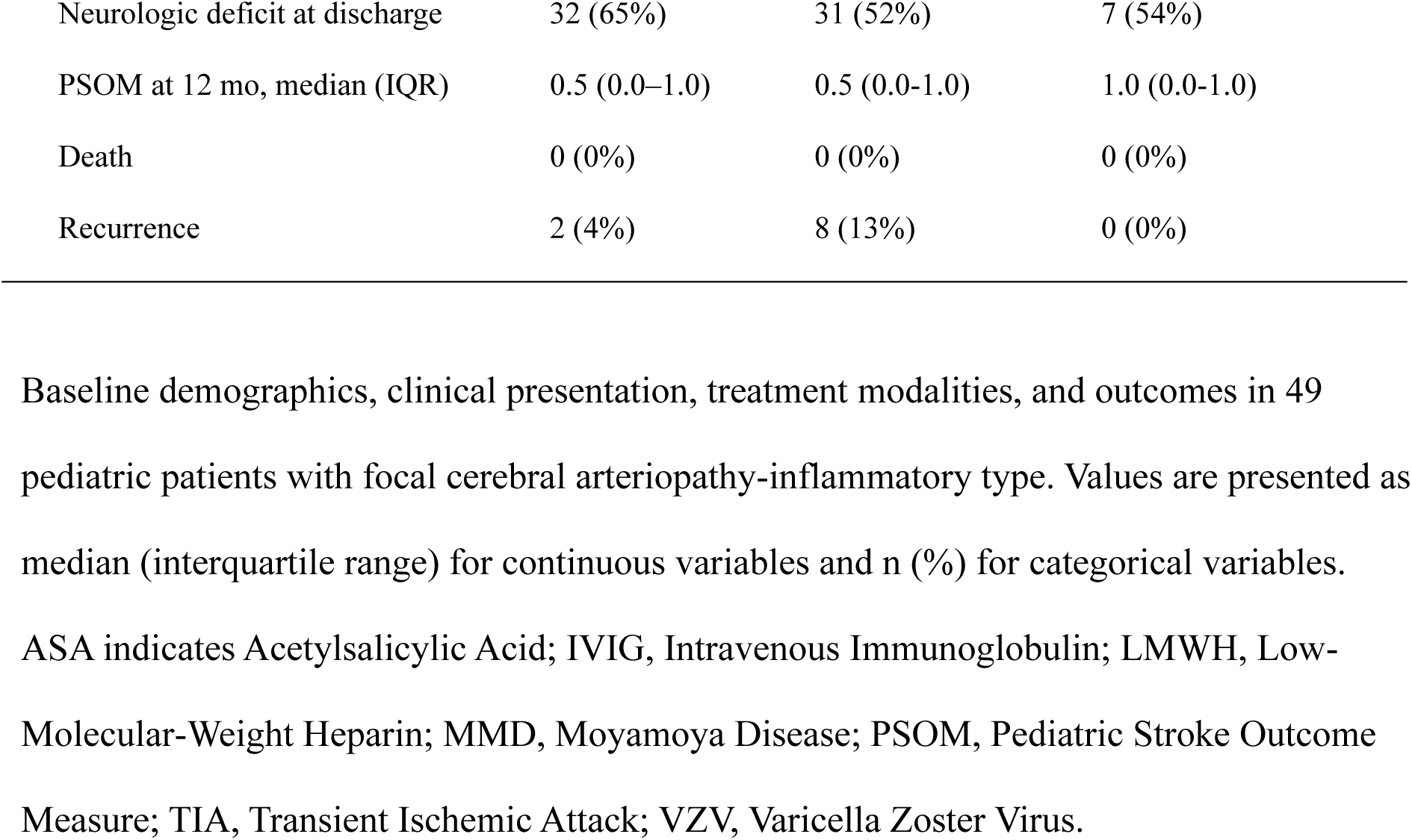
Baseline Characteristics of Pediatric Arteriopathy Subtypes.

### Neuroimaging Outcomes and FCASS Dynamics in FCA-i

Of the 49 patients with FCA-i, 41 (84%) with anterior circulation involvement were included in neuroimaging analyses, while posterior cases were excluded. Serial neuroimaging revealed a median of 4.0 brain MRIs per FCA-i patient (IQR, 2.0–6.0), with initial MRI performed within 1 day of symptom onset. The median neuroimaging follow-up duration was 2.1 years (IQR, 0.7–4.0). Initial infarct burden, assessed by modified pediatric Alberta Stroke Program Early CT Score (modASPECTS), was modest (median 3.0; IQR, 2.0–4.0), and baseline FCASS was low (median 2.0; IQR, 2.0–4.0), indicating mild initial arteriopathy severity (Table 2).

**Table 2.** Neuroimaging Characteristics and Focal Cerebral Arteriopathy Severity Score.

| Neuroimaging outcomes | FCA-i (n=41) | uMMD (n=17) |
| --- | --- | --- |
| Brain MRIs performed per patient | 4.0 (2.0–6.0) | 6.0 (4.0–7.0) |
| Time of initial MRI following stroke symptoms, d | 1.0 (0.0–1.0) | 1.0 (0.0–1.0) |
| Neuroimaging follow-up, y | 2.1 (0.7–4.6) | 5.0 (3.7–11.3) |
| Initial acute modASPECTS | 3.0 (2.0–4.0) | 3.0 (2.0–4.8) |
| Initial FCASS | 2.0 (2.0–4.0) | 6.0 (4.0–8.5) |
| Worst FCASS | 4.0 (3.0–6.0) | 8.0 (7.0–12.0) |
| Maximum worsening FCASS | 4.0 (2.0–5.0) | 2.5 (2.0–5.5) |
| Time to worst FCASS, m | 2.0 (1.0–5.0) | 2.7 (0.6–9.5) |
| Maximum improvement FCASS | 2.0 (0.0–4.0) | 0.0 (0.0–2.0) |
| Time to lowest FCASS, m | 11.0 (5.0–16.0) | 6.6 (0.7–32.0) |
| Final FCASS | 3.0 (1.0–5.0) | 8.0 (5.0–12.0) |
| FCASS change initial to final | 0.0 (–2.0–2.0) | 2.0 (0.0–2.0) |

Serial magnetic resonance imaging findings and Focal Cerebral Arteriopathy Severity Score evolution in patients with focal cerebral arteriopathy-inflammatory type (FCA-i) and unilateral moyamoya disease (uMMD). Analyses were limited to 41 FCA-i patients with anterior circulation involvement; posterior circulation cases (n = 8) were excluded. For uMMD, inclusion was based on unilateral involvement with anterior circulation involvement, confirmed by angiography, resulting in 17 patients. Values are presented as median (interquartile range). FCASS indicates Focal Cerebral Arteriopathy Severity Score; modASPECTS, Modified Pediatric Alberta Stroke Program Early CT Score; MRI, Magnetic Resonance Imaging..

### FCASS Validation: Correlation with Infarct Burden and Temporal Evolution

A significant positive correlation was observed between initial modASPECTS and FCASS (Spearman ρ=0.42, *P*=0.0069), validating the association between vascular lesion severity and acute infarct burden (Figure 1). Across follow-up imaging, FCASS demonstrated a characteristic temporal evolution, showing an early peak (median maximum FCASS = 4.0; IQR, 2.0–5.0) followed by gradual improvement to a final median of 3.0 (IQR, 1.0–5.0). The time to maximum FCASS was 2.0 months (IQR, 1.0–5.0), and the time to lowest FCASS was 11.0 months (IQR, 5.0–16.0), indicating a monophasic disease course with partial vascular recovery (Figure 2).

**Figure 1.**
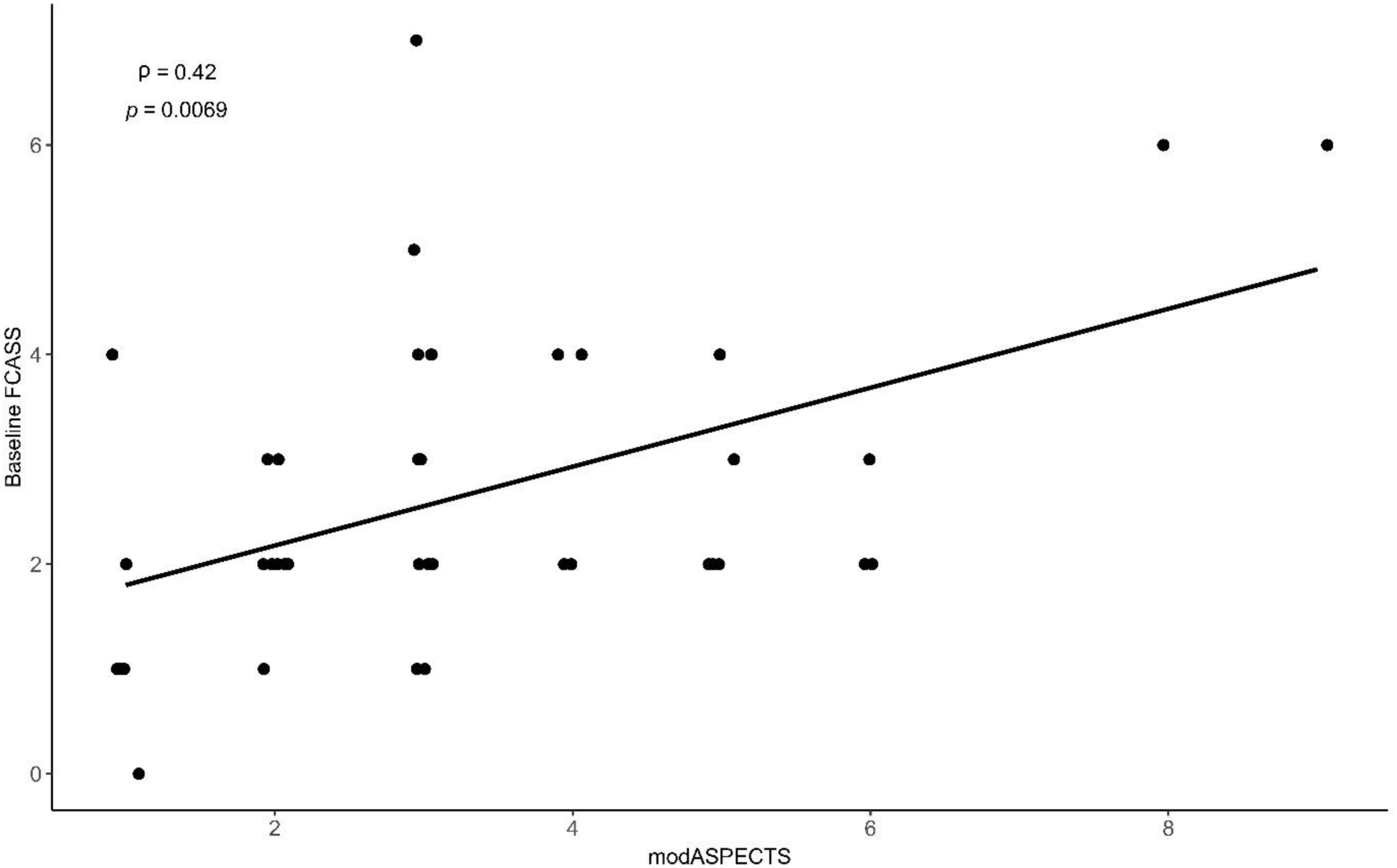
Correlation Between Baseline Infarct Burden and Focal Cerebral Arteriopathy Severity Score. Scatter plot demonstrating correlation between baseline modified pediatric Alberta Stroke Program Early CT Score and Focal Cerebral Arteriopathy Severity Score in patients with focal cerebral arteriopathy-inflammatory type (n=41). Spearman rank correlation was used for statistical analysis (ρ=0.42, *P*=0.0069). Linear regression line with 95% confidence interval is shown. modASPECTS indicates modified pediatric Alberta Stroke Program Early CT Score; FCASS, Focal Cerebral Arteriopathy Severity Score.

**Figure 2.**
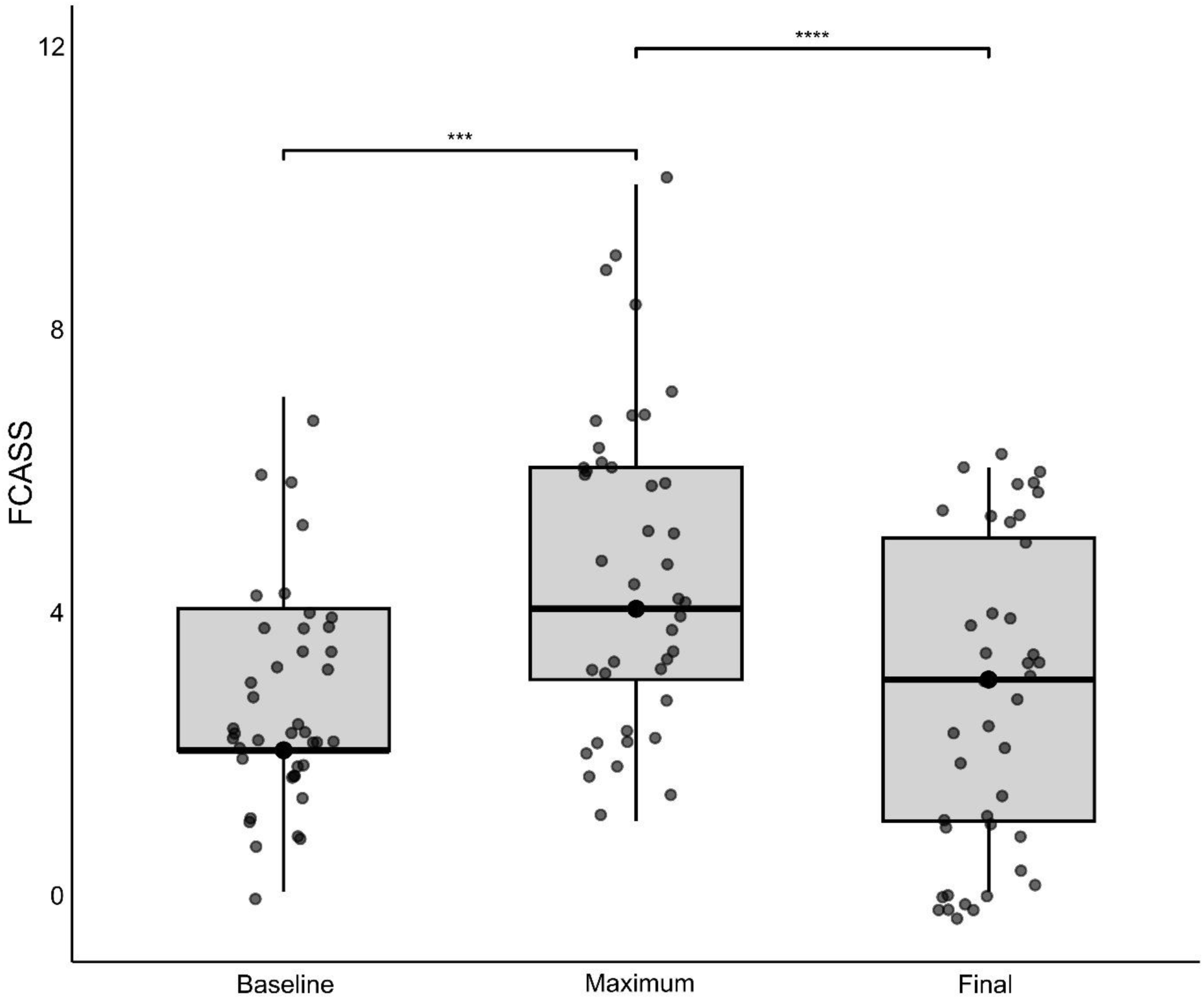
Focal Cerebral Arteriopathy Severity Score Distribution at Baseline, Maximum, and Final Follow-up. Comparison of Focal Cerebral Arteriopathy Severity Score at baseline, maximum, and final follow-up timepoints in patients with focal cerebral arteriopathy-inflammatory type (n=41). Box plots display median (center line), interquartile range (box), and 1.5× interquartile range (whiskers); individual data points are overlaid. Statistical analysis was performed using Wilcoxon signed-rank tests with Bonferroni correction for multiple comparisons. Significant worsening was observed from baseline to maximum scores (\*\*\**P*<0.001), followed by significant improvement at final follow-up compared with maximum (\*\*\*\**P*<0.001). FCASS indicates Focal Cerebral Arteriopathy Severity Score; IQR, interquartile range.

### Longitudinal FCASS Trajectories

Among FCA-i patients with serial neuroimaging, 21 (51%) demonstrated early progression of arteriopathy followed by stabilization or improvement (Figure 3A). Population-level smoothing analysis using locally weighted scatterplot smoothing (LOWESS, span = 0.75) revealed a consistent temporal trajectory characterized by an early peak within several months after onset, followed by gradual improvement beginning around 6 months post-stroke (Figure 3B). Analysis of FCASS changes (ΔFCASS) by time intervals showed the most significant increase within 3 months post-onset, followed by significant decrease in the 6- to 12-month interval, with sustained stability through 24 months. Bonferroni-corrected comparisons demonstrated statistically significant differences between early (0–3 months) and late (6–12 and 12–24 months) intervals (*P*<0.05 for all comparisons) (Figure 4).

**Figure 3.**
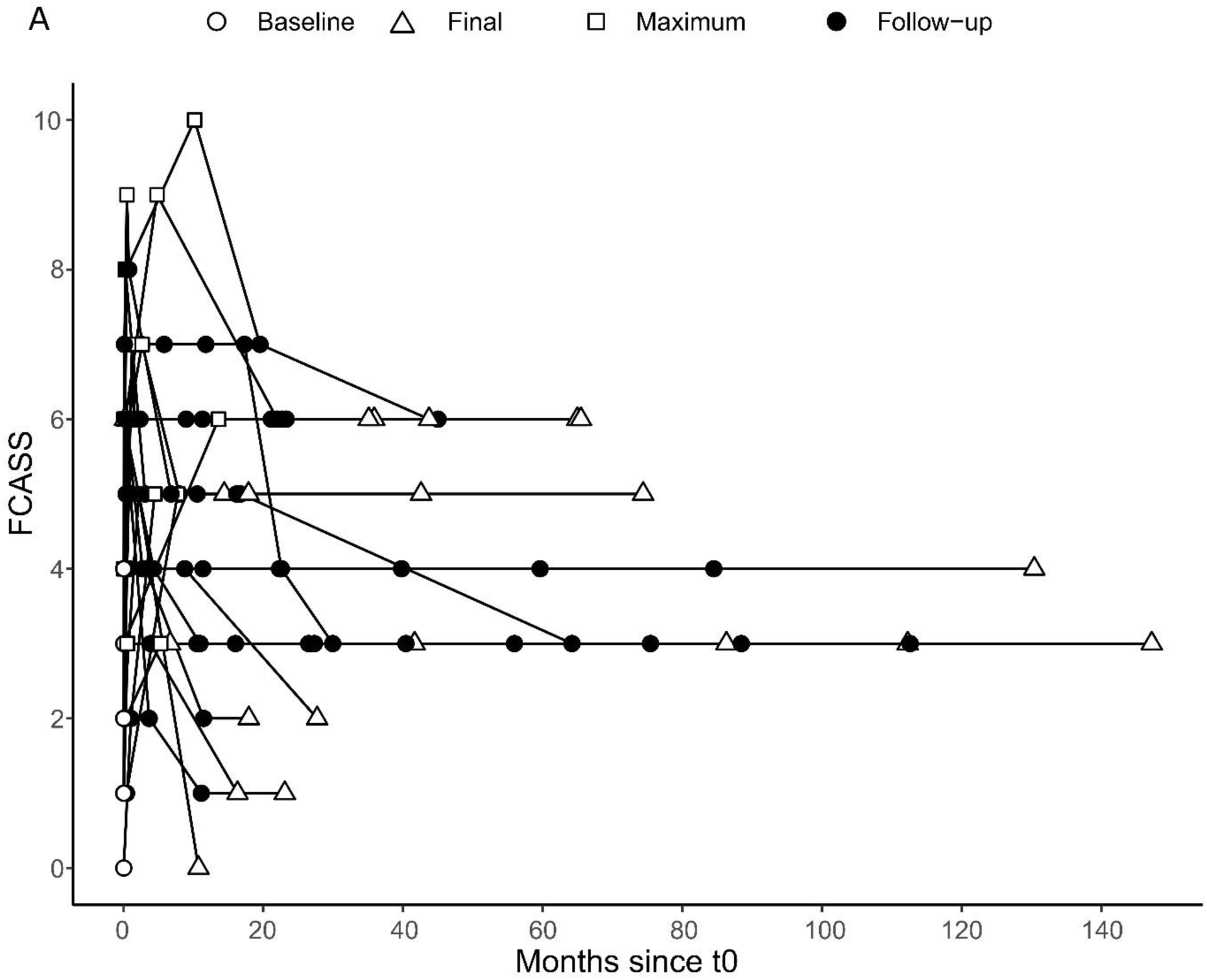
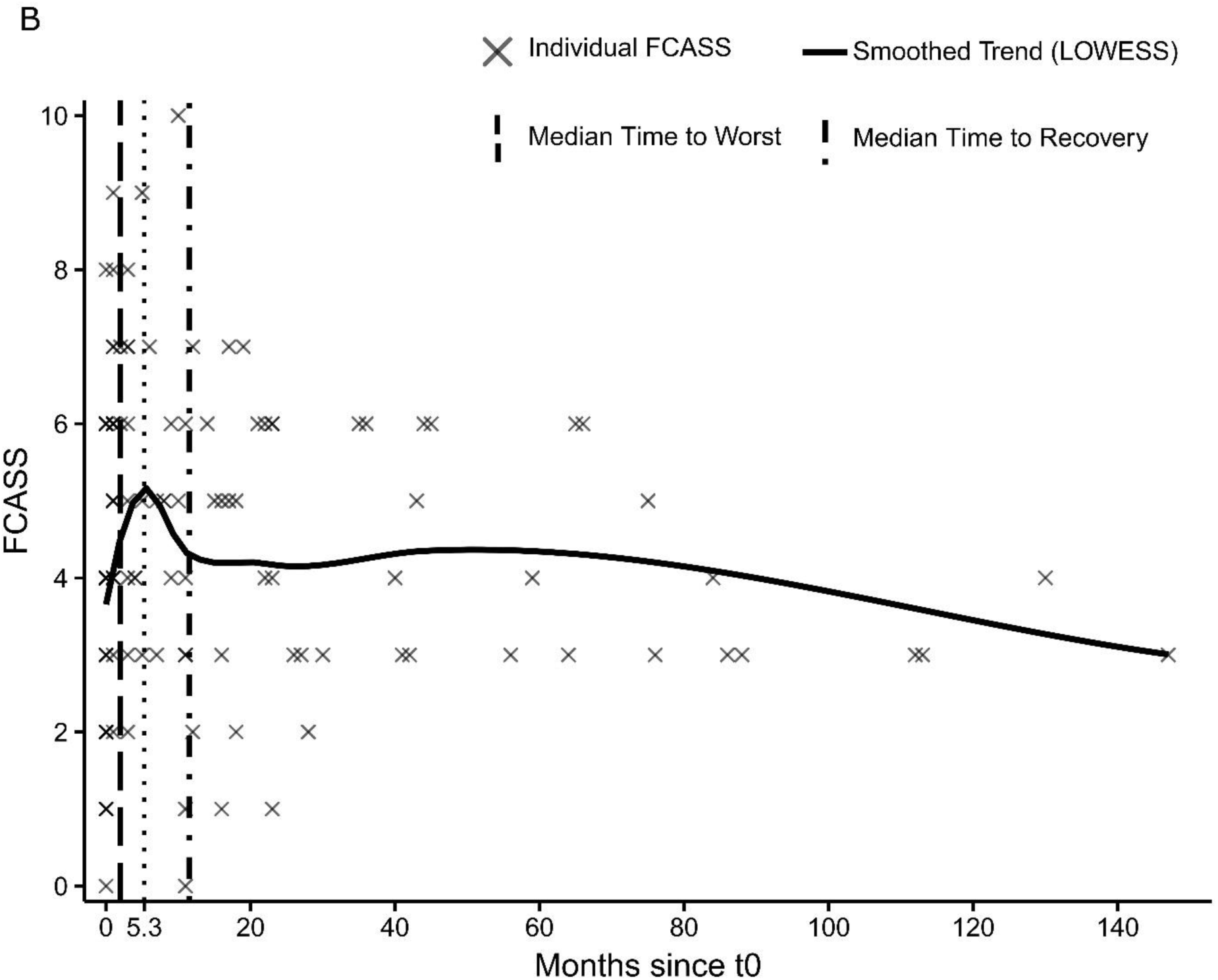
Individual and Population-Level Focal Cerebral Arteriopathy Severity Score Trajectories. Panel A, Individual longitudinal trajectories of Focal Cerebral Arteriopathy Severity Score in patients with worsening arteriopathy (n=21). Each line represents a unique patient, with FCASS values plotted over time since baseline. Most patients exhibited early worsening of FCASS within the first few months, followed by stabilization or improvement, highlighting the monophasic nature of focal cerebral arteriopathy-inflammatory type and the importance of early intensive vascular imaging follow-up. Panel B, Longitudinal trend of FCASS scores in all patients with focal cerebral arteriopathy-inflammatory type. Individual FCASS measurements (gray crosses) are shown along with a smoothed trend line using locally weighted scatterplot smoothing (LOWESS; solid black line). The vertical dashed line represents the median time to peak FCASS, and the dotted line indicates the median time to recovery. The overall pattern demonstrates rapid arteriopathy worsening in the early months, followed by gradual improvement within approximately 12 months. FCASS indicates Focal Cerebral Arteriopathy Severity Score; LOWESS, Locally Weighted Scatterplot Smoothing.

**Figure 4.**
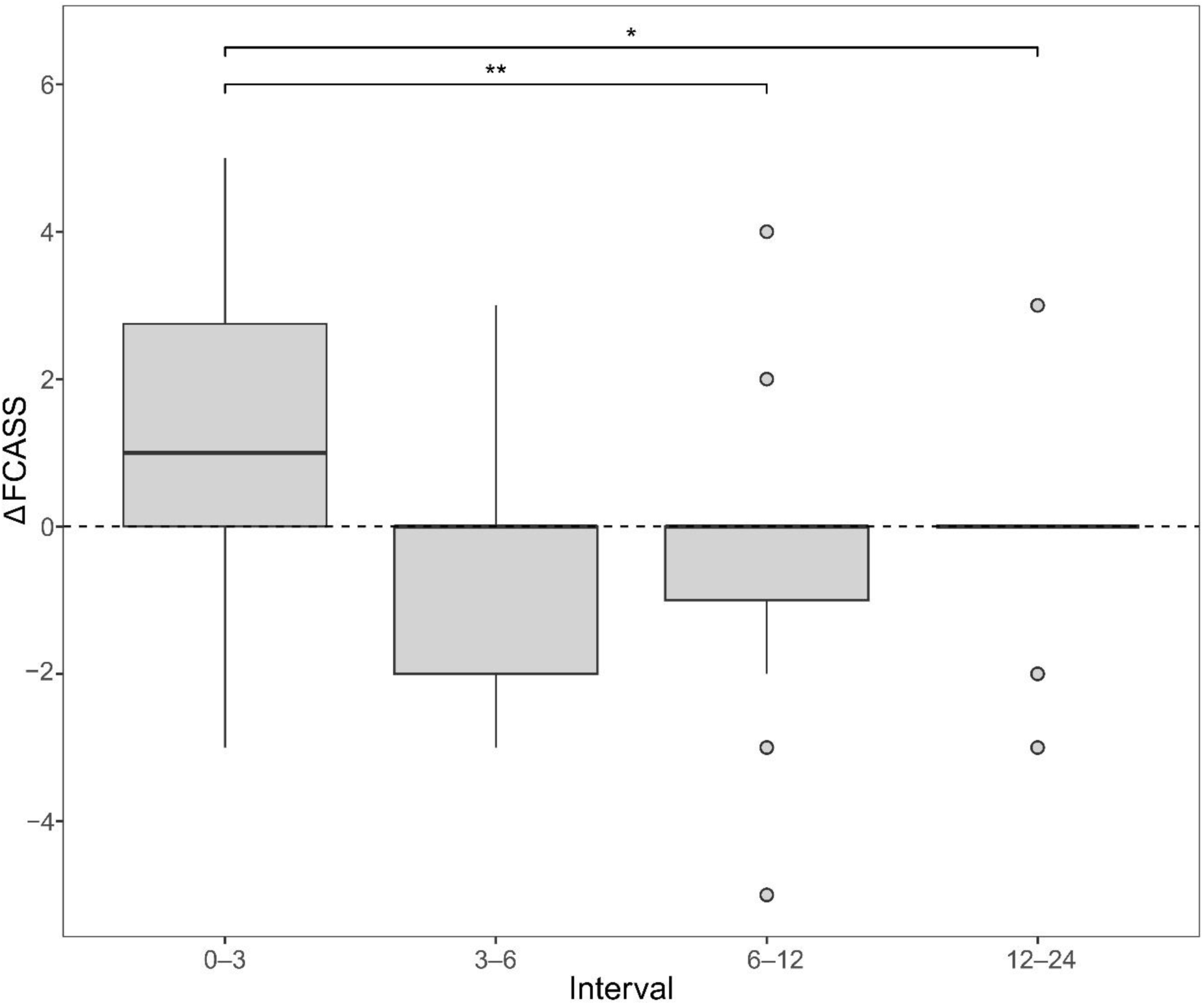
Focal Cerebral Arteriopathy Severity Score Changes by Time Intervals. Box plots showing Focal Cerebral Arteriopathy Severity Score changes across predefined time intervals in patients with focal cerebral arteriopathy-inflammatory type who demonstrated radiographic worsening (n=21). Positive values indicate worsening, whereas negative values indicate improvement. Significant changes were observed between early and late intervals (0–3 vs 6–12 months, *P*<0.01; 0–3 vs 12–24 months, *P*<0.05). Comparisons were performed using Wilcoxon signed-rank tests with Bonferroni correction for multiple comparisons. Boxes represent the interquartile range (IQR), center lines indicate the median, and whiskers denote 1.5×IQR. FCASS indicates Focal Cerebral Arteriopathy Severity Score; IQR, interquartile range.

### Comparative Analysis: FCA-i vs Unilateral MMD

Both FCA-i and unilateral MMD patients were observed over sufficiently long neuroimaging follow-up durations—2.1 years (IQR, 0.7–4.0) for FCA-i and 5.0 years (IQR, 3.7–11.3) for unilateral MMD—providing a reliable basis for longitudinal comparison. Early arteriopathy worsening occurred in both groups within the first few months after stroke onset, with median times to peak FCASS of 2.0 months (IQR, 1.0–5.0) in FCA-i and 2.7 months (IQR, 0.6–9.5) in unilateral MMD. Despite this initial similarity, their subsequent trajectories diverged markedly (Table 2).

Across all timepoints—baseline, maximum, and final—unilateral MMD patients exhibited consistently higher FCASS scores than those with FCA-i (Figure 5A). Median baseline FCASS in unilateral MMD was 6.0 (IQR, 4.0–8.5) compared with 2.0 (IQR, 2.0–4.0) in FCA-i (*P*<0.001); this difference remained significant at both maximum FCASS (8.0 [7.0–12.0] vs. 4.0 [3.0–6.0], *P*<0.001) and final follow-up (8.0 [5.0–12.0] vs. 3.0 [1.0–5.0], *P*<0.001). Notably, only FCA-i demonstrated radiographic improvement over time, with a median FCASS decrease of 2.0 points (IQR, 0.0–4.0) and lowest values observed around 11 months post-onset. In contrast, unilateral MMD showed no measurable improvement (ΔFCASS = 0.0 [0.0–2.0]), consistent with the progressive, non-reversible nature of MMD vasculopathy.

**Figure 5.**
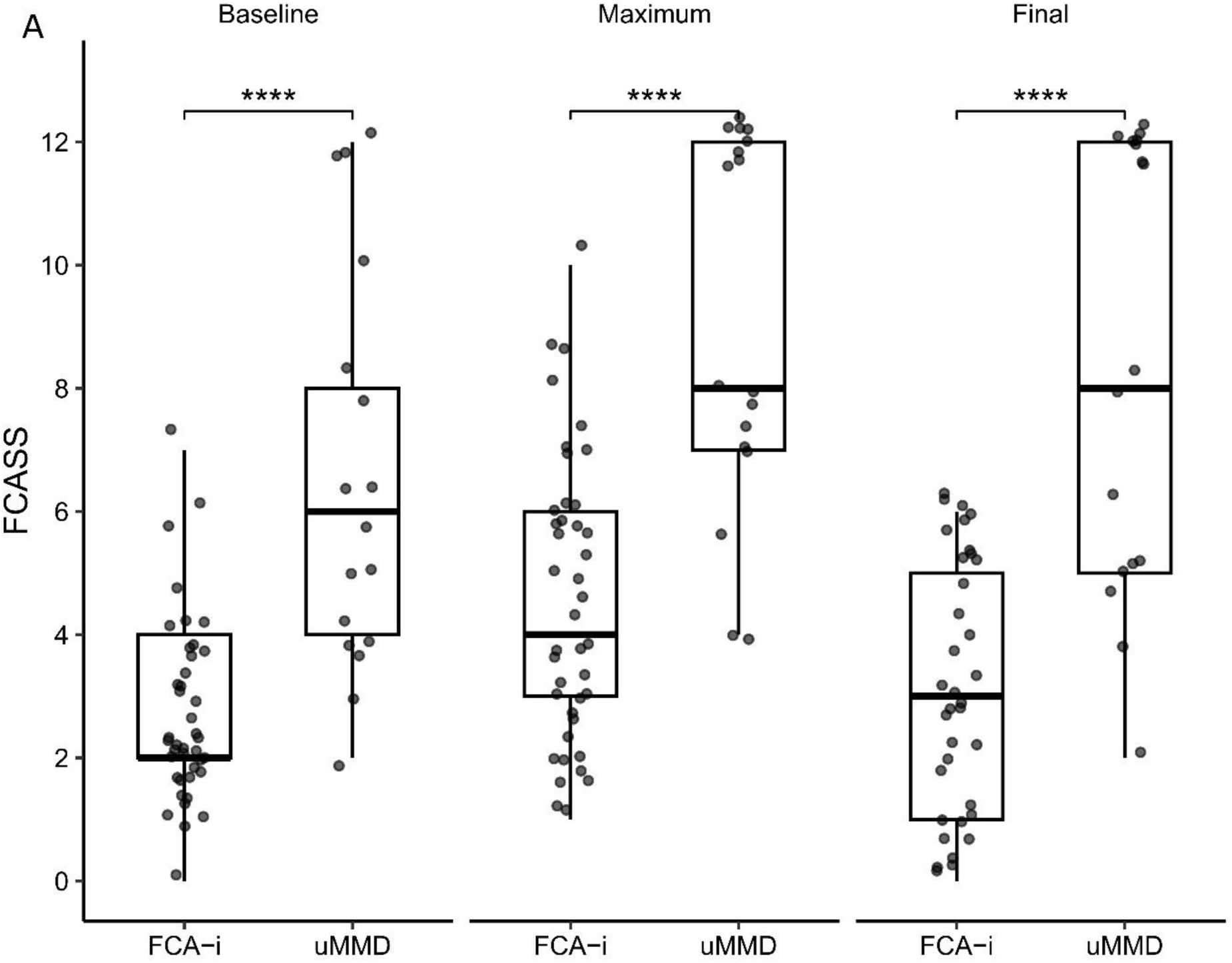
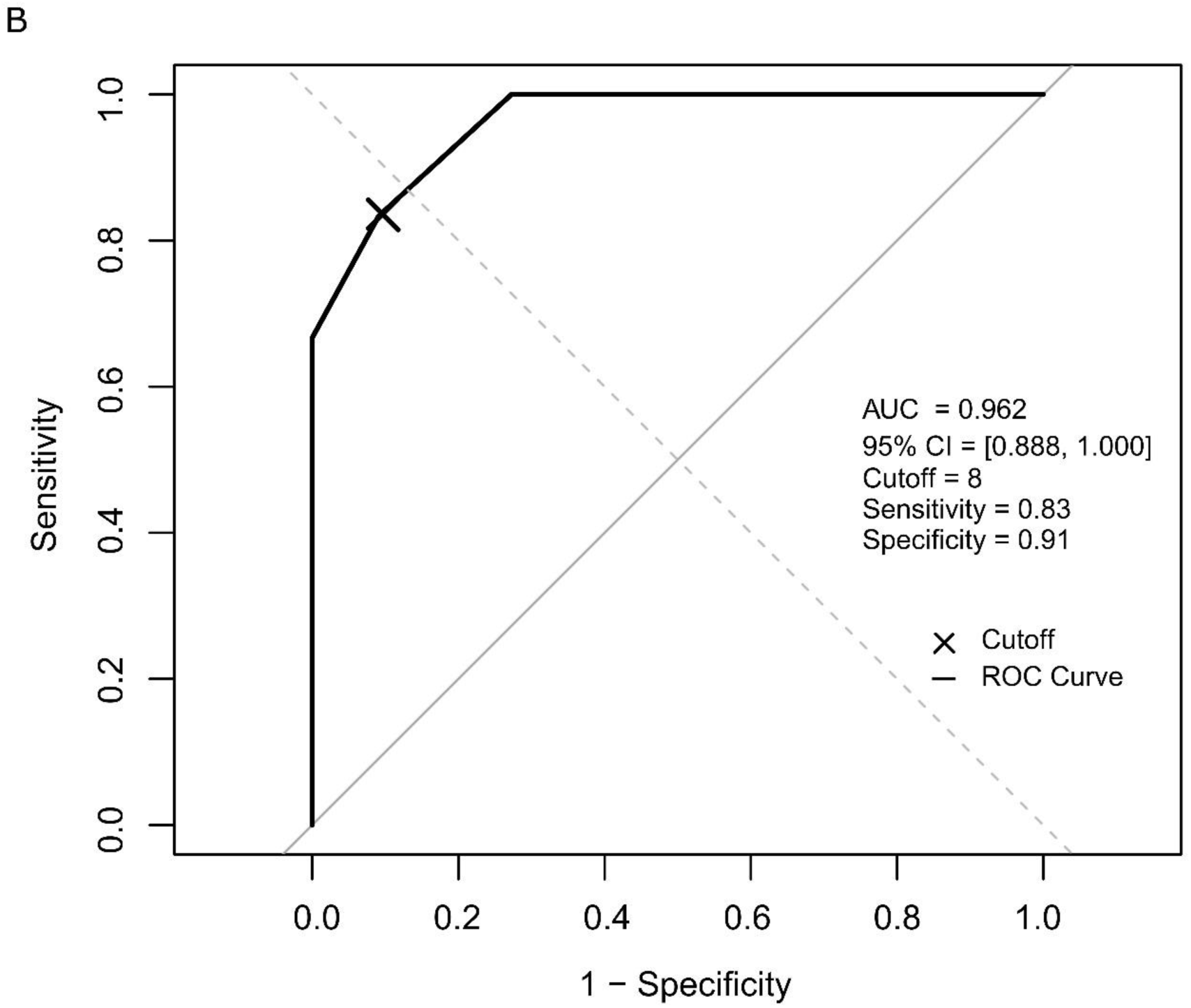
Comparative Analysis of Focal Cerebral Arteriopathy Severity Score Between FCA-i and Unilateral Moyamoya Disease. Panel A, Comparison of Focal Cerebral Arteriopathy Severity Score at baseline, maximum, and final timepoints between focal cerebral arteriopathy-inflammatory type (n=41) and unilateral moyamoya disease (n=17). Unilateral MMD patients showed significantly higher FCASS values at all timepoints (\*\*\*\**P*<0.0001, Mann–Whitney *U* test). Box plots display the median (center line), interquartile range (box), and 1.5× interquartile range (whiskers), with individual data points overlaid. Panel B, Receiver operating characteristic (ROC) curve analysis evaluating baseline FCASS for predicting contralateral progression in unilateral MMD (n=17). An optimal cutoff of ≥8.0 yielded an area under the curve (AUC) of 0.962 (95% CI, 0.888–1.000), with a sensitivity of 0.83 and specificity of 0.91. FCA-i indicates focal cerebral arteriopathy-inflammatory type; uMMD, unilateral moyamoya disease; FCASS, Focal Cerebral Arteriopathy Severity Score; ROC, receiver operating characteristic; AUC, area under the curve.

### FCASS as Predictor of Contralateral Progression

Among 17 patients with unilateral MMD, 6 patients (35%) developed contralateral progression during follow-up. Baseline FCASS values were significantly higher in patients with progression (10.0 ± 2.5) than in those without (4.6 ± 1.6, *P*=0.002). In univariate logistic regression, baseline FCASS was associated with contralateral progression both as a continuous variable (odds ratio = 3.06, 95% CI 1.46–16.0, *P*=0.049) and when dichotomized at ≥8 vs <8 (odds ratio = 50.0, 95% CI 3.83–2038.0, *P*=0.010).

Receiver operating characteristic (ROC) analysis identified a baseline FCASS cutoff of ≥8.0 as the optimal threshold for predicting contralateral progression, yielding an area under the curve of 0.962 (95% CI 0.888–1.000; sensitivity, 0.83; specificity, 0.91) (Figure 5B). These findings support baseline FCASS ≥8 as a potential imaging biomarker for early risk stratification in unilateral MMD.

## DISCUSSION

We validated the clinical utility and temporal dynamics of the FCASS in a Korean pediatric cohort with FCA-i and contrasted its neuroimaging trajectory with unilateral MMD. Through longitudinal analysis of serial brain MRAs, our findings reinforce FCASS as a dynamic biomarker of disease severity and progression in FCA-i, and demonstrate its potential role in differentiating monophasic inflammatory arteriopathy from progressive arteriopathies in regions with high MMD prevalence.

Previous studies have established FCASS as a reproducible measure correlating with infarct burden and clinical outcomes in FCA across North American and European cohorts.^15–17^ However, its performance has not been evaluated in East Asian populations, where the high endemic prevalence of MMD presents unique diagnostic challenges.^7,18^ In our cohort, FCASS correlated significantly with modASPECTS at baseline (ρ=0.42, *P*=0.0069) and exhibited the characteristic monophasic pattern of early worsening followed by gradual recovery, consistent with an inflammatory arteriopathy. These findings validate the external applicability of FCASS in an ethnically distinct population and support its integration into diagnostic algorithms for FCA-i across diverse regions.

Our study’s strength lies in dense temporal sampling with a median of four MRAs per patient over a 2.1-year follow-up period. This imaging density enabled detailed characterization of the longitudinal trajectories of arteriopathy severity. Consistent with previous studies, FCASS peaked early, typically within 2–3 months after symptom onset, followed by a gradual decline that stabilized by 12 months in most patients.^10,12,18^

Subgroup analysis of patients with radiographic worsening (n=21) revealed statistically significant deterioration within the first 3 months, followed by meaningful recovery in the 6–12-month interval. These temporal patterns support short-interval imaging during the acute and subacute phases and suggest a practical follow-up schedule: MRAs at diagnosis, 1–2 months (to capture peak disease), and 6–12 months (to assess recovery). Beyond 12 months, follow-up imaging may be individualized according to clinical status or new neurological symptoms. This recommendation aligns with recent consensus that serial vascular imaging remains essential for monitoring, given limitations of inflammatory serum biomarkers and emerging vessel wall imaging modalities.^9,25,26^

Distinguishing FCA-i from early unilateral MMD remains a major diagnostic challenge, particularly in East Asia where MMD prevalence is significantly higher than in Europe or North America.^7,18,27^ Both entities can present with unilateral large-vessel stenosis in anterior circulation and similar clinical symptoms such as hemiparesis and basal ganglia infarction. Early-stage MMD may lack well-developed collateralization, further complicating differentiation on initial imaging. This diagnostic ambiguity is reflected in inter-rater reliability studies showing only moderate reproducibility for FCA-i (κ=0.40–0.49).^14^ Our study highlights this challenge by comparing FCASS trajectories between FCA-i and unilateral MMD. Although both groups showed early worsening, only FCA-i demonstrated subsequent radiographic recovery. This monophasic course supports the concept of FCA-i as a self-limited inflammatory arteriopathy, in contrast to the progressive, steno-occlusive vasculopathy characteristic of MMD.

While FCASS was originally developed to quantify arteriopathy severity in FCA, its anatomic basis allows extension to other unilateral large-vessel arteriopathies.^15,28^ In our exploratory analysis, unilateral MMD patients exhibited consistently higher FCASS scores across all timepoints, suggesting more extensive and persistent vascular involvement compared with FCA-i. Notably, ROC analysis revealed that baseline FCASS ≥8.0 provided high discrimination for predicting contralateral progression in unilateral MMD (area under the curve=0.962). In patients with baseline FCASS ≥8, the involved segments commonly included the distal ICA and proximal A1 or M1. These findings suggest that high FCASS values reflect more extensive arteriopathy affecting the terminal ICA region, which may explain the higher likelihood of contralateral progression observed in this subgroup.

Given that contralateral progression remains a significant concern in unilateral MMD, with rates ranging from 30%–50%, early identification of high-risk patients could inform surveillance imaging timing and potential surgical intervention.^19–21,29,30^ Nevertheless, FCASS application in MMD should be interpreted cautiously, as it was not originally developed for this disease. Prospective multicenter studies are needed to determine whether FCASS can reliably predict MMD progression across diverse populations.

Based on our findings, we propose a tiered follow-up imaging framework for pediatric patients with unilateral arteriopathy: initial diagnosis with FCASS assessment and modASPECTS evaluation; early follow-up (1–2 months) to identify peak severity; intermediate follow-up (6–12 months) to assess recovery; and extended follow-up (12+ months) for patients with persistent stenosis or high progression risk (FCASS ≥8.0). This approach integrates vascular severity scoring with known natural history to inform evidence-based imaging intervals.

Our study has several limitations. The retrospective single-center design introduces potential selection bias and may limit generalizability. Variable MRI follow-up intervals between patients may affect precise estimation of FCASS peak timing. Subtle contralateral abnormalities may have been underrecognized at baseline, potentially confounding progression prediction analysis. While FCASS was applied to unilateral MMD exploratorily, formal validation remains necessary before routine use. Finally, inflammatory markers were inconsistently available and excluded from multivariable models; their potential adjunctive role warrants future prospective evaluation.

## CONCLUSIONS

This study validates FCASS as a dynamic imaging biomarker for FCA-i and demonstrates its utility in distinguishing monophasic inflammatory arteriopathy from progressive MMD. This distinction is particularly critical in East Asian populations where MMD prevalence is high. FCASS-based monitoring enables identification of disease progression patterns and recovery trajectories, informing evidence-based follow-up strategies and serving as a prognostic tool for contralateral progression in unilateral MMD.

Future research should focus on prospective multicenter validation of FCASS, integration with vessel wall imaging and inflammatory biomarkers, and randomized trials evaluating corticosteroid efficacy in focal cerebral arteriopathy.^25,31–33^ These evidence-based approaches will enhance diagnostic precision and therapeutic decision-making in pediatric AIS.

### Nonstandard Abbreviations and Acronyms

FCASS: Focal Cerebral Arteriopathy Severity Score
FCA: Focal Cerebral Arteriopathy
FCA-i: Focal cerebral arteriopathy–inflammatory type
CASCADE: Childhood AIS Standardized Classification and Diagnostic Evaluation
AIS: Arterial Ischemic Stroke
MMD: Moyamoya Disease

## Data Availability

The data supporting this study are available from the corresponding author upon reasonable request.

## Notes

### Competing Interest Statement

The authors have declared no competing interest.

### Funding Statement

No external funding was received.

### Author Declarations

The study was approved by Seoul National Hospital's institutional review board, which waived the need for written informed consent based on minimal risk (IRB No. H-2508-155-1670).

